# Cancer and risk of COVID-19 through a general community survey

**DOI:** 10.1101/2020.05.20.20103762

**Authors:** Karla A. Lee, Wenjie Ma, Daniel R. Sikavi, Jonathan Wolf, Claire J. Steves, Tim D. Spector, Andrew T. Chan, on behalf of the COPE consortium

**Affiliations:** Department of Twin Research and Genetic Epidemiology, King’s College London, London, U.K.; Clinical and Translational Epidemiology Unit and Division of Gastroenterology, Massachusetts General Hospital and Harvard Medical School. Boston, MA, USA.; Department of Medicine, Massachusetts General Hospital and Harvard Medical School, Boston, MA, USA.; Zoe Global Limited. London, U.K.

## Abstract

**Background:** Data are limited on the risk of coronavirus disease 2019 (COVID-19) among individuals with cancer and whether cancer-related therapy exacerbates this risk.

**Methods:** We evaluated the risk for COVID-19 among patients living with cancer compared to the general community and whether cancer-related treatments influence this risk. Data were collected from the COVID Symptom Study smartphone application since March 24, 2020 (United Kingdom), March 29 (U.S.), and April 29, 2020 (Sweden) through May 8, 2020. We used multivariate-adjusted odds ratios (aORs) of a positive COVID-19 test as well as predicted COVID-19 infection using a validated symptom model.

**Results:** Among 23,266 participants with cancer and 1,784,293 without cancer, we documented 10,404 reports of a positive COVID-19 test. Compared to participants without cancer, those living with cancer had 62% increased risk of a positive COVID-19 test (95% CI: 1.37-1.91). Among patients with cancer, current treatment with chemotherapy/immunotherapy was associated with a nearly 2.5-fold increased risk of a positive test (aOR: 2.42; 95% CI: 1.81-3.25). The association between cancer and COVID-19 positivity was stronger among participants >65 years (aOR: 1.93; 95% CI: 1.51-2.46) compared to younger participants (aOR: 1.32; 95% CI: 1.06-1.64; P_interaction_ <0.001); and among amles (aOR: 1.71; 95% CI: 1.36-2.15) compared to females (aOR; 95% CI: 1.14-1.79; P_interaction_ =0.02).

**Conclusions:** Individuals with cancer had a significantly increased risk of infection compared to the general community. Those treated with chemotherapy or immunotherapy were particularly at-risk of infection.

**Trial Registration:** ClinicalTrials.gov NCT04331509

## Introduction

Individuals with cancer may be at higher risk for coronavirus disease 2019 (COVID-19). However, data are limited largely to small studies conducted among hospitalized patients.

## Methods

We recruited individuals from the general population in the United Kingdom, United States, and Sweden using The COVID Symptom Study, a freely available smartphone application developed by Zoe Global Ltd. in collaboration with the Massachusetts General Hospital and King’s College London offering a guided interface to report a range of baseline demographic information and comorbidities as previously reported.^1^ Participants are encouraged to use the application daily to report symptoms and COVID-19 testing results. We queried if individuals were living with cancer (yes/no) and if they were on chemotherapy or immunotherapy (yes/no) beginning on March 29, 2020. We employed multivariable logistic regression models to examine the association between cancer and risk of a positive COVID-19 test (COVID-19+). We separately analyzed the risk associated with chemotherapy/immunotherapy for COVID-19+ among individuals with cancer. Two-sided *p*-values<0.05 were considered statistically significant. All analyses were performed using R 3.6.1 (Vienna, Austria).

## Results

Through May 8, 2020, 1,807,559 participants provided demographic and longitudinal symptom and testing information. Compared to individuals without cancer, those with cancer were older, more frequently male, and more commonly overweight/obese, among other comorbidities (**Table 1**). They were more likely to use several common medications and have health problems requiring them to stay at home. Among 23,266 individuals with cancer and 1,784,293 without cancer, we documented 155 and 10,249 reports of a positive COVID-19+ test, respectively (**Table 2**). Compared to individuals without cancer, those with cancer had a 60% increased risk of COVID-19+ (adjusted odds ratio (aOR): 1.60; 95% confidence interval (CI): 1.36-1.88). The association between cancer and COVID-19+ was stronger among participants >65 years (aOR: 1.93; 95%CI: 1.51-2.46) compared to younger participants (aOR: 1.32; 95%CI: 1.06-1.64; P_interaction_<0.001); and among males (aOR: 1.71; 95%CI: 1.36-2.15) compared to females (aOR: 1.43; 95%CI: 1.14-1.79; P_interaction_=0.02). Chemotherapy or immunotherapy was associated with a 2-fold increased risk of COVID-19+ (aOR: 2.22; 95%CI: 1.68-2.94). An increased risk of hospitalization due to COVID-19 was associated with a cancer diagnosis (aOR: 2.47; 95%CI: 2.22-2.76) and chemotherapy/immunotherapy (aOR: 4.16; 95%CI: 2.50-4.95). Using a validated symptom-based prediction model for COVID-19,^2^ the aOR for predicted COVID-19 was 1.32 (95% CI:1.22-1.42) for those with cancer and 1.55 (95%CI: 1.33-1.79) for those on chemotherapy/immunotherapy.

**Table 1.** Baseline characteristics of participants according to cancer history and chemotherapy or immunotherapy.

|  | Cancer |  | Chemotherapy/immunotherapy |  |
| --- | --- | --- | --- | --- |
|  | No<br>(n=1,784,293) | Yes<br>(n=23,266) | No<br>(n=1,802,655) | Yes<br>(n=4,904) |
| **Country (%)** |  |  |  |  |
| U.K. | 81.6 | 77.1 | 81.5 | 75.1 |
| U.S. | 11.8 | 18.3 | 11.9 | 19.7 |
| Sweden | 6.6 | 4.7 | 6.6 | 5.2 |
| **Age group (%)** |  |  |  |  |
| <25 | 15.5 | 0.9 | 15.3 | 1.8 |
| 25-34 | 14.1 | 1.0 | 14.0 | 1.7 |
| 35-44 | 17.0 | 3.9 | 16.9 | 5.9 |
| 45-54 | 18.9 | 10.7 | 18.8 | 14.8 |
| 55-64 | 17.3 | 23.1 | 17.4 | 23.4 |
| >=65 | 17.2 | 60.3 | 17.6 | 52.5 |
| **Male sex (%)** | 42.7 | 55.2 | 42.9 | 45.8 |
| **Ethnicity (%)** |  |  |  |  |
| Hispanic | 5.9 | 3.5 | 5.9 | 4.4 |
| Non-Hispanic | 90.2 | 93.3 | 90.2 | 91.7 |
| Prefer not to say | 3.9 | 3.3 | 3.9 | 3.9 |
| **Race (%)** |  |  |  |  |
| White | 93.6 | 95.6 | 93.7 | 94.9 |
| Black | 1.4 | 1.0 | 1.4 | 1.1 |
| Asian | 2.5 | 1.7 | 2.5 | 2.0 |
| Other | 2.0 | 1.2 | 2.0 | 1.4 |
| Prefer not to say | 0.4 | 0.4 | 0.4 | 0.5 |
| **Body mass index group (%)** |  |  |  |  |
| <18.5 | 6.5 | 3.3 | 6.4 | 4.1 |
| 18.5-24.9 | 40.4 | 37.0 | 40.3 | 38.7 |
| 25-29.9 | 31.1 | 36.5 | 31.2 | 33.8 |
| >=30 | 22.0 | 23.2 | 22.0 | 23.3 |
| **Comorbidities (%)** |  |  |  |  |
| Diabetes | 4.0 | 10.2 | 4.1 | 10.3 |
| Heart disease | 3.4 | 12.6 | 3.5 | 10.6 |
| Lung disease | 12.1 | 17.1 | 12.1 | 18.4 |
| Kidney disease | 0.8 | 4.5 | 0.9 | 4.8 |
| **Smoking status (%)** |  |  |  |  |
| Never | 70.8 | 61.6 | 70.7 | 63.7 |
| Past | 20.2 | 33.2 | 20.4 | 31.4 |
| Current | 9.0 | 5.3 | 8.9 | 5.0 |
| **Limited mobility (%)<sup>a</sup>** | 7.1 | 40.9 | 7.4 | 64.1 |
| **Medication use (%)** |  |  |  |  |
| Immunosuppressants<sup>b</sup> | 3.5 | 16.3 | 3.5 | 43.7 |
| ACE inhibitor | 7.3 | 17.1 | 7.4 | 15.4 |
| Aspirin | 4.8 | 16.3 | 4.9 | 17.5 |
| NSAIDs | 7.4 | 10.8 | 7.5 | 10.8 |
| **Interaction with individuals with COVID-19 (%)** |  |  |  |  |
| No | 87.0 | 93.2 | 87.1 | 94.5 |
| Yes, suspected | 9.5 | 4.8 | 9.4 | 3.8 |
| Yes, documented | 3.5 | 2.0 | 3.5 | 1.7 |
| **Frontline healthcare worker (%)** | 7.2 | 2.8 | 7.1 | 2.1 |

Abbreviations: ACE (angiotensin converting enzyme), NSAIDs (non-steroidal anti-inflammatory drugs). Proportions are calculated based on the total number of participants with available data. History of cancer, uses of aspirin and NSAIDs, and smoking status have been queried since launch in the U.S. and Sweden and since 3/29/2020 in the U.K.
<sup>a</sup> Immunosuppressant medications including steroids, methotrexate, biologics were asked.
<sup>b</sup> Limited mobility was asked as “In general, do you have any health problems that require you to stay at home?”

**Table 2.**
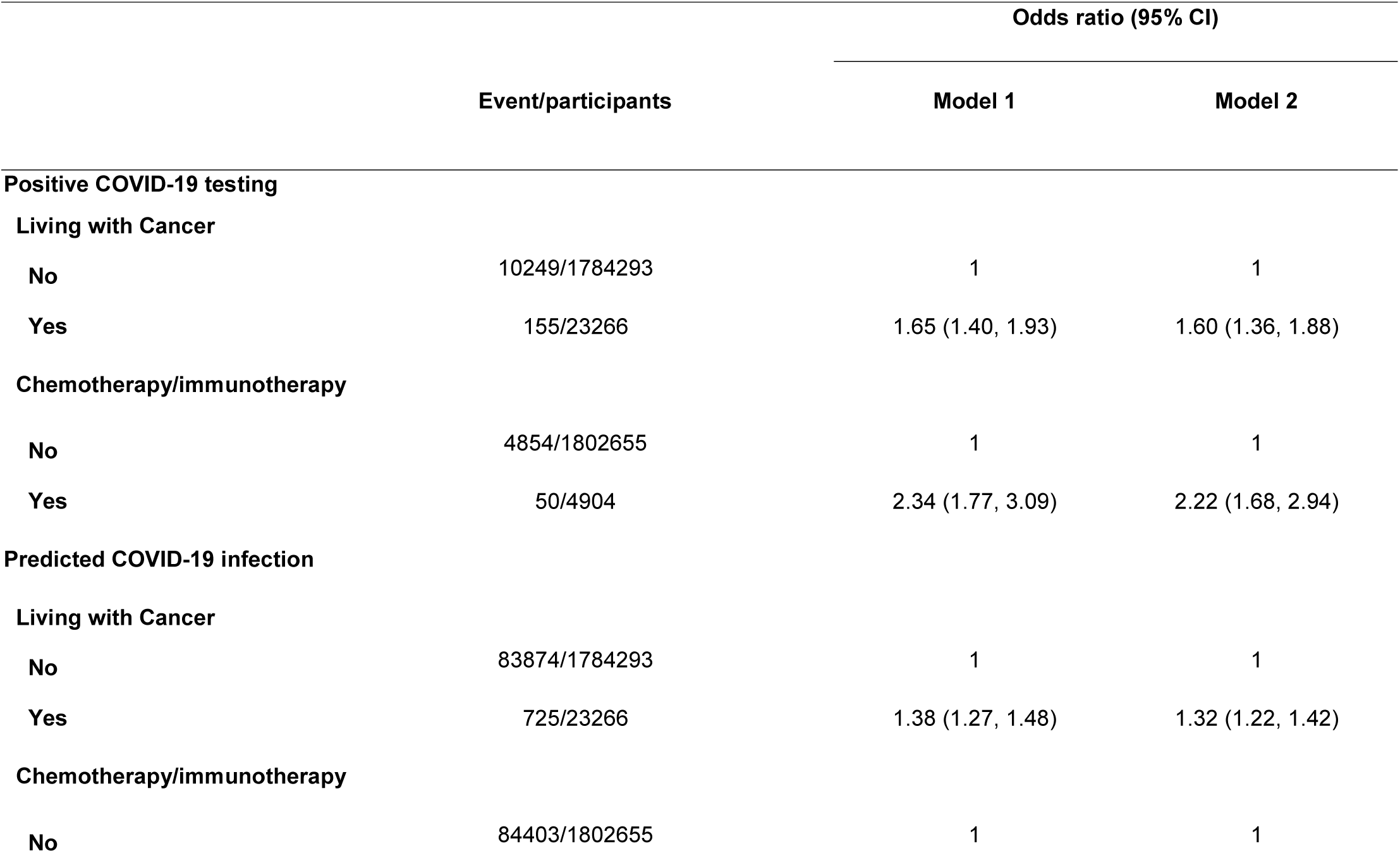
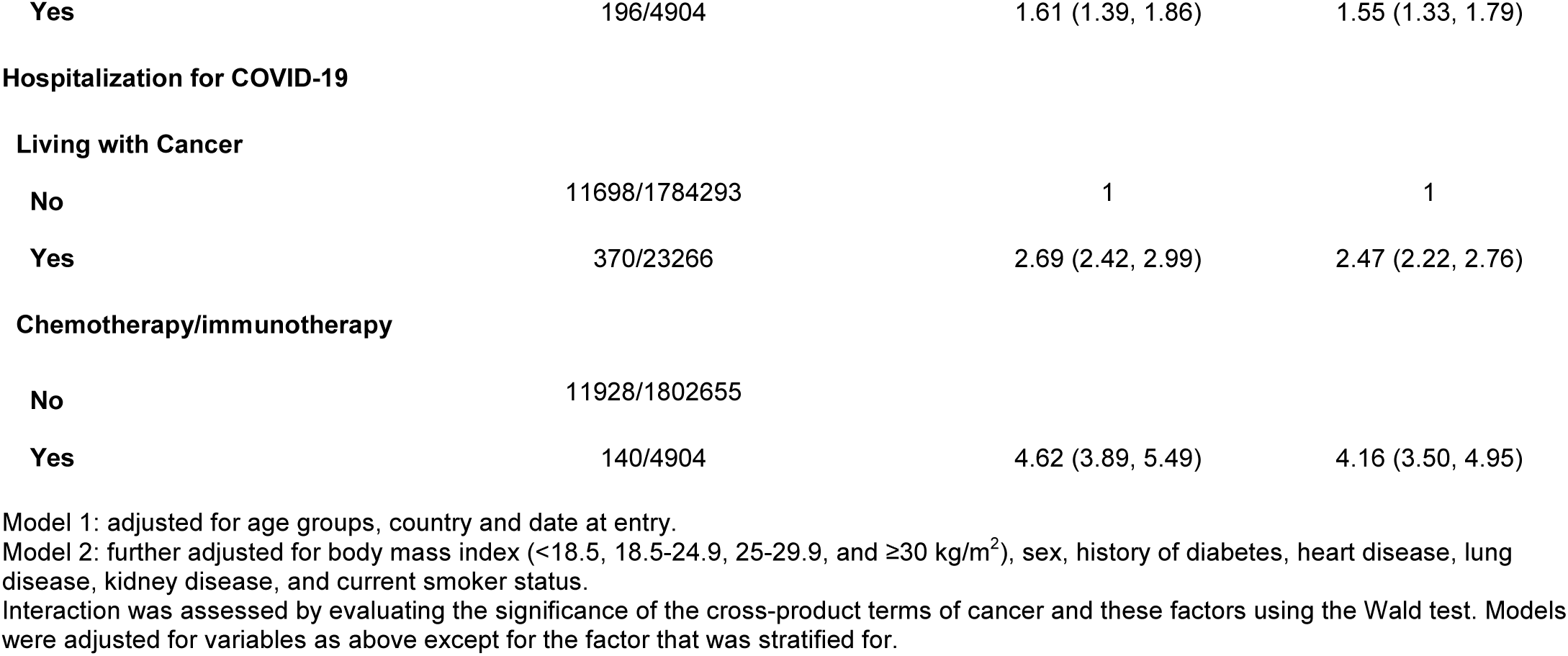
Associations between cancer history, chemotherapy/immunotherapy and risk of COVID-19.

## Discussion

Among >1.8 million participants, we found that individuals living with cancer had a 60% increased risk of COVID-19+ or hospitalization with COVID-19, with greater risks for older individuals or those receiving anti-cancer therapies. Prior studies have shown that individuals with cancer comprise a disproportionate share of poorer COVID-19 outcomes,^3-6^ including death. However, these studies had small sample sizes and are largely based on hospitalized patients, capturing the most severe cases. Individuals living with cancer also tend to be older with greater comorbidities that predispose to hospitalization and adverse events. Our results from a large, community-based sample support that incidence of infection, including milder disease with more limited symptoms, is also higher in individuals with cancer. Our study was limited by the use of self-reported information collected from individuals who used smartphone devices. Covid-19 testing was not based on uniform screening. However, the current shortage of PCR-based testing kits in both the U.K. and the U.S. does not make such an approach feasible. We had limited data on specific tumor types and treatment regimens. We are planning future studies collecting more detailed information from individuals with cancer with linkage to other data sources.

## Data Availability

Data collected in the app are being shared with other health researchers through the NHS-funded Health Data Research UK (HDRUK)/SAIL consortium, housed in the UK Secure e-Research Platform (UKSeRP) in Swansea. Anonymized data collected by the symptom tracker app can be shared with bonafide researchers via HDRUK, provided the request is made according to their protocols and is in the public interest (see https://healthdatagateway.org/detail/9b604483-9cdc-41b2-b82c-14ee3dd705f6). US investigators are encouraged to coordinate data requests through the COPE Consortium (www.monganinstitute.org/cope-consortium). Data updates can be found at https://covid.joinzoe.com.

## Funding

Zoe provided in kind support for all aspects of building, running and supporting the tracking app and service to all users worldwide. King’s College of London investigators (KAL, CJS, TDS) were supported by the Wellcome Trust and EPSRC (WT212904/Z/18/Z, WT203148/Z/16/Z, T213038/Z/18/Z), the NIHR GSTT/KCL Biomedical Research Centre, MRC/BHF (MR/M016560/1), and the Alzheimer’s Society (AS-JF-17-011). ATC is the Stuart and Suzanne Steele MGH Research Scholar and Stand Up to Cancer scientist. An Evergrande COVID-19 Response Fund Award through the Massachusetts Consortium on Pathogen Readiness (MassCPR) and Mark and Lisa Schwartz also supported MGH investigators (WM, DRS, ATC). Support was also received from the Swedish Foundation for Strategic Research (LUDC-IRC 15-0067), the Swedish Heart-Lung Foundation and the Swedish research Council.

## Acknowledgements

We would like to thank the more than 3 million contributing citizen scientists who have downloaded the COVID Symptom Study, including participants of cohort studies within the COronavirus Pandemic Epidemiology (COPE) Consortium. We thank the investigators including Long H. Nguyen, David A. Drew, Amit D. Joshi, Chuan-Guo Guo, and Chun-Han Lo from the MGH Clinical and Translational Epidemiology Unit (CTEU); Sebastien Ourselin, Carole H. Sudre, Mary Ni Lochlainn, Thomas Varsavsky, Mark Graham, M.Jorge Cardoso, Cristina Menni, Marc Modat, Ruth Bowyer, Maxim B Freidin, Benjamin Murray, Alessia Visconti and Veronique Bataille from King’s College London; Paul W. Franks and Maria Gomez from Lund University Diabetes Center; and Tove Fall from Uppsala University for their assistance with data collection, analysis and writing.

We also thank the investigators of the cohort studies enrolled in the COPE Consortium; the MGH CTEU Clinical Research Coordination team; Sophie Papa, Paul Nathan and Heather Shaw for development of cancer-related questions; the staff of Zoe Global Ltd for providing technical support for the app; Stand Up to Cancer for their assistance in media and social media outreach.

## Ethics

In the UK, the App Ethics has been approved by KCL ethics Committee REMAS ID 18210, review reference LRS-19/20-18210 and all subscribers provided consent. In Sweden, ethics approval for the study was provided by the central ethics committee (DNR 2020-01803).

## Conflicts of Interest

JW is an employee of Zoe Global Ltd. TDS is a consultant to Zoe Global Ltd. ATC previously served as an investigator on a clinical trial of diet and lifestyle using a separate mobile application that was supported by Zoe Global Ltd. Other authors have no conflict of interest to declare.

## Notes

### Clinical Trial

NCT04331509

